# Bilateral jaw thrust usage is neither universally prevalent during laryngeal mask airway placement nor universally prevalent during anesthesia mask ventilation before laryngoscopy for endotracheal tube placement: A single anesthesia worksite observation

**DOI:** 10.1101/2023.11.22.23298890

**Authors:** Deepak Gupta, Kaya Chakrabortty, Mohamed Ismaeil

## Abstract

**Background:** While placing two hands on the mask during manual or mechanical ventilation on unsecured airways, providers are not always providing bilateral jaw thrust for maintaining airway patency especially when focusing only on ensuring a good mask seal.

**Objectives:** To assess a single anesthesia worksite real-world prevalence of bilateral jaw thrust usage, it was looked through either (a) during laryngeal mask airway placement, or (b) during anesthesia mask ventilation before laryngoscopy for endotracheal tube placement.

**Materials and Methods:** Over a period of two months, anesthesia care of patients was respectively observed in terms of either (a) various strategies involved during laryngeal mask airway placement in its successful placement, or (b) various strategies involved during anesthesia mask ventilation in successful ventilation before laryngoscopy for endotracheal tube placement.

**Results:** During the two-month period, a total of 213 patients were observed. Among the small number of observed very first attempts at laryngeal mask airway placement (n=13), bilateral jaw thrust was only observed in 69% attempts. Among the total observations before the very first attempts at laryngoscopy for endotracheal tube placement (n=200), 26% were devoid of any attempts at anesthesia mask ventilation before laryngoscopy. Among the remaining observations (n=148), anesthesia mask ventilations were attempted before the very first attempts at laryngoscopy for endotracheal tube placement but bilateral jaw thrust was only observed in 37% attempts.

**Conclusion:** During laryngeal mask airway placement and during anesthesia mask ventilation before laryngoscopy for endotracheal tube placement, bilateral jaw thrust usage was not universally prevalent at the observed single anesthesia worksite.

## Introduction

It has been believed that for maintaining airway patency [1], bilateral jaw thrust can be a major skill in the armamentarium of anesthesia care providers and other healthcare providers who are providing ventilatory support, manually or mechanically, to patients’ airways which have yet not been secured with in-situ advanced airway devices like endotracheal tube and laryngeal mask airway. It has also been observed anecdotally that bilateral jaw thrust can facilitate laryngeal mask airway placement. Ironically, while placing two hands on the mask during manual or mechanical ventilation on unsecured airways, providers are not always providing bilateral jaw thrust especially when focusing only on ensuring a good mask seal [2]. Therefore, it was deemed a necessary quality assurance (QA) initiative to explore the prevalence of bilateral jaw thrust usage among patients who were scheduled to receive anesthesia care via airway secured with laryngeal mask airway or endotracheal tube.

Henceforth, two QA projects were designed to assess a single anesthesia worksite’s real-world prevalence of bilateral jaw thrust usage either (a) during laryngeal mask airway placement, or (b) during anesthesia mask ventilation before laryngoscopy for endotracheal tube placement.

## Materials and Methods

Institutional review board deemed these QA projects as non-human participant research. These QA projects were concurrently conducted over a period of two months wherein anesthesia care of patients was respectively observed in terms of either (a) various strategies involved during laryngeal mask airway placement in its successful placement, or (b) various strategies involved during anesthesia mask ventilation in patients’ successful ventilation before laryngoscopy for endotracheal tube placement.

Various strategies observed during the very first attempt at laryngeal mask airway placement were:

- Mandibular depression [3]
- Bilateral jaw thrust [4-6]
- Tongue depression [7]
- Finger guidance inside mouth [8]
- Crossed-scissor maneuver [9]

Confirmed visualization of capnograph [10-12] after the very first attempt at laryngeal mask airway placement deemed it as a successful placement.

Various strategies observed during anesthesia mask ventilation before the very first attempt at laryngoscopy for endotracheal tube placement were:

- Manual bag ventilation
- Mechanical ventilation
- No ventilation
- One hand one provider on anesthesia mask
- Two hands one provider on anesthesia mask
- More than two hands meaning more than one provider on anesthesia mask
- Bilateral jaw thrust
- Oropharyngeal airway
- Nasopharyngeal airway
- One provider doing everything
- Two providers doing it together
- Three or more providers doing it as team

Confirmed visualization of capnograph during anesthesia mask ventilation before the very first attempt at laryngoscopy for endotracheal tube placement deemed it as a successful ventilation.

## Results

During the two-month period, a total of 213 patients were observed with only 13 patients observed during laryngeal mask airway placement while 200 patients observed during anesthesia mask ventilation before laryngoscopy for endotracheal tube placement. This suggested that for every laryngeal mask airway placement to secure airway, there were at least 15 endotracheal tube placements to secure airway at the observed single anesthesia worksite thus suggesting laryngeal mask airway as not the preferred/feasible advanced airway device at the observed single anesthesia worksite.

Among the small number of observed very first attempts at laryngeal mask airway placement (n=13), bilateral jaw thrust was only observed in 69% attempts while mandibular depression, tongue depression, finger guidance inside mouth and crossed-scissor maneuver observed in 85% attempts, 54% attempts, 69% attempts and 15% attempts respectively suggested that more than one among the five observed strategies were being concurrently utilized during laryngeal mask airway placements which were thence observed to be 85% successful in the very first attempts.

Among the total observations before the very first attempts at laryngoscopy for endotracheal tube placement (n=200), 26% were devoid of any attempts at anesthesia mask ventilation before laryngoscopy. Among the remaining observations (n=148), anesthesia mask ventilations were attempted before the very first attempts at laryngoscopy for endotracheal tube placement (n=148) and their strategies have been detailed in Table 1. As compared to manual ventilation with anesthesia reservoir bags (n=137), mechanical ventilation with anesthesia workstations’ ventilators (n=11) was not the preferred mode of pre-laryngoscopy anesthesia mask ventilation at the observed single anesthesia worksite with at least 12 manual ventilations observed pre-laryngoscopy for every mechanical ventilation observed pre-laryngoscopy. Still mechanical ventilation, while universally utilizing bilateral jaw thrust, was observed to be 100% successful in anesthesia mask ventilation as compared to success rates varying from 79%-91% among manual ventilations depending on the variable number of providers and their hands involved in maintaining mask seal with or without bilateral jaw thrust during manual ventilations with anesthesia reservoir bags. Although 19% attempts at pre-laryngoscopy anesthesia mask ventilation (n=148) involved oropharyngeal airway usage as well, none of the observed pre-laryngoscopy anesthesia mask ventilations involved nasopharyngeal airway usage thus suggesting a potential avoidance of nasopharyngeal airway usage at the observed single anesthesia worksite.

**Table 1:**
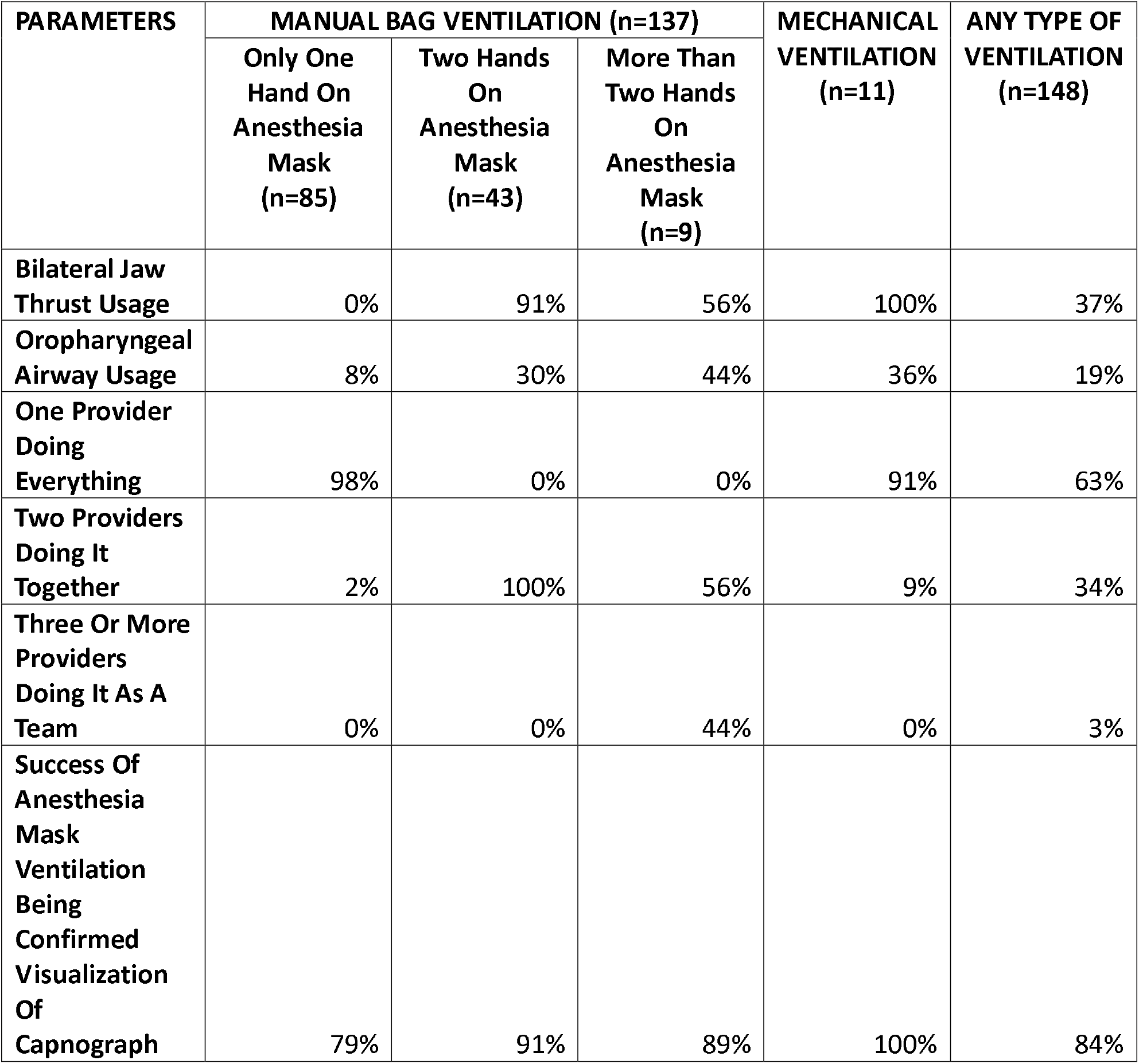
Percentage About Prevalence Of Pre-Laryngoscopy Anesthesia Mask Ventilation Parameters

## Discussion

At the observed single anesthesia worksite, the key findings of these concurrently completed QA projects were the following: (a) laryngeal mask airway placement [13] was observed to be not the preferred advanced airway device among providers, most likely due to not being the feasible choice for securing airway as guided by patient-related risk factors; (b) more than one among the five observed strategies were being concurrently utilized during laryngeal mask airway placements, most likely due to none of the strategies in itself being an effective solo strategy to achieve success in laryngeal mask airway placement; (c) 26% laryngoscopy for endotracheal tube placement were devoid of any attempts at anesthesia mask ventilation before laryngoscopy, most likely due to patient-related risk factors warranting rapid sequence induction/intubation [14-15] with easy access to video-laryngoscopy concurrently soaring providers’ confidence level in safely securing airway without first attempting and checking the adequacy of anesthesia mask ventilation; (d) mechanical ventilation with anesthesia workstations’ ventilators [16-17] was not the preferred mode of pre-laryngoscopy anesthesia mask ventilation, most likely due to providers’ historically attuned comfort in manual ventilation with anesthesia reservoir bags during pre-laryngoscopy anesthesia mask ventilation; (e) nasopharyngeal airway [18-19] was never utilized during pre-laryngoscopy anesthesia mask ventilation, most likely due to unknown nasal and nasopharyngeal patency risking mucosal bleeding therein with unanticipated nasopharyngeal airway placement in unprepared nasal cavity; and (f) bilateral jaw thrust usage [20-22] was neither universally prevalent during laryngeal mask airway placement nor universally prevalent during anesthesia mask ventilation before laryngoscopy for endotracheal tube placement, most likely due to patient-related risk factors warranting providers’ forced and unforced choices to avoid bilateral jaw thrust usage.

These QA projects had some limitations. The total number of observations could not be extended beyond 213 due to research team’s logistics thus preventing the key findings of these QA projects from convincingly making or breaking the case for bilateral jaw thrust and other strategies utilized either during laryngeal mask airway placement or during anesthesia mask ventilation before laryngoscopy for endotracheal tube placement. Moreover, during the observation periods, the providers could have unanticipatedly changed their routinely employed strategies during laryngeal mask airway placement and during anesthesia mask ventilation before laryngoscopy for endotracheal tube placement. Finally, patient-related risk factors guiding providers’ personal reasons for choosing particular strategies during laryngeal mask airway placement and during anesthesia mask ventilation before laryngoscopy for endotracheal tube placement could not be explored due to these QA projects being non-human participant research.

## Conclusion

Summarily, during laryngeal mask airway placement and during anesthesia mask ventilation before laryngoscopy for endotracheal tube placement at the observed single anesthesia worksite, bilateral jaw thrust usage was not universally prevalent. Moreover, more than quarter of very first attempts at laryngoscopy for endotracheal tube placement did not have adequacy of successful anesthesia mask ventilation checked pre-laryngoscopy. Finally, nasopharyngeal airway usage and mechanical ventilation with anesthesia workstations’ ventilators were underutilized during pre-laryngoscopy anesthesia mask ventilation.

## Supporting information

Non-Human Participant Reseach Per IRB-LMA

Non-Human Participant Reseach Per IRB-ETT

## Data Availability

All data produced in the present work are contained in the manuscript

## Notes

### Competing Interest Statement

The authors have declared no competing interest.

### Funding Statement

This study did not receive any funding

### Author Declarations

Wayne State University: IRB Administration Office determined that these QA projects did not meet the definition of Human Participant Research subject to IRB oversight and review

