## Supplementary material for "Bilateral jaw thrust usage is neither universally prevalent during laryngeal mask airway placement nor universally prevalent during anesthesia mask ventilation before laryngoscopy for endotracheal tube placement: A single anesthesia worksite observation": Non-Human Participant Reseach Per IRB-ETT

**IRB Administration Office**
87 E. Canfield, Second Floor
Detroit, MI 48201
www.research.wayne.edu/irb

### Notice of IRB Administrative Determination: Non-Human Participant Research (HPR)

**To:** Deepak Gupta, MD
**From:** Heather Park-May
**Date:** May 17, 2023
**Re:** Variable Strategies In Anesthesia Mask Ventilation By Our Anesthesia Provider Teams

| | | | | |
|---|---|---|---|---|
| Project Title: | Variable Strategies In Anesthesia Mask Ventilation By Our Anesthesia Provider Teams | | | |
| Name of person conducting the project: | Deepak Gupta | Title: | MD | Date: 5/12/2023 |
| Email Address: | | Phone: | 313-966-3004 | |
| Status: **Select all that apply** | ☑ Wayne State Faculty<br>☐ WSU Graduate Student<br>☐ WSU Undergraduate Student<br>☐ Resident/Fellow/Trainee<br>☐ Other: | ☑ DMC Staff<br>☑ Karmanos Staff<br>☐ J. D. Dingell VAMC Staff | | |
| Division or College: | Harper University Hospital | Department: | Anesthesiology | |
| Campus Address: | Box No 162, 3990 John R, Detroit, MI-48201 | | | |
| Faculty Sponsor/ Supervisor for this Project: | Name:<br>☐ I do not have a Faculty Sponsor/Supervisor | Title:<br>Phone: | | |
| Form completed by: | Deepak Gupta | E-mail: | | |
| **Check ALL that apply:** | ☑ Behavioral, social, education, non-medical study<br>☑ Medical study | | | |

| | |
|---|---|
| **7.** | Describe how the data will be obtained (e.g. survey, interview, observation, testing, review of existing records, etc.):<br>Observation by anesthesia researchers not involved as a team member in the anesthesia care to record the variations in strategies employed by anesthesia teams when ventilating by anesthesia mask in their patients presenting for procedures under general endotracheal tube anesthesia. Primarily the students on research rotation will follow anesthesia teams planning to perform general endotracheal tube anesthesia for anesthesia care wherein they will record percentage of successful anesthesia mask ventilations wherein anesthesia teams will be variably using one or more of the following strategies based on their personal preference and clinical experience with the strategies:<br>(a) bag ventilation, and/or<br>(b) mechanical ventilation, and/or<br>(c) no ventilation<br>plus<br>(a) one hand one provider on mask, and/or<br>(b) two hands one provider on mask, and/or<br>(c) more than two hands meaning more than one provider on mask, and/or<br>(d) bilateral jaw-thrust, and/or<br>(e) oral airway, and/or<br>(f) nasopharyngeal airway<br>plus<br>(a) one person doing everything<br>(b) two persons doing it together<br>(c) three or more persons doing it as team |

#### Section C: Description of the Project:

**Provide a description of the project with enough detail for the determination.**

| | |
|---|---|
| **8.** | Describe the purpose of this project:<br>The purpose of this project is to record the variations in strategies employed by anesthesia team when ventilating by anesthesia mask in their patients presenting for procedures under general endotracheal tube anesthesia at Harper University Hospital and to quantify percentage of successful anesthesia mask ventilations as corresponding to strategies employed by our anesthesia teams at Harper University Hospital |

| | |
|---|---|
| **9.** | Describe the objectives or aims for the project:<br>Observation by anesthesia researchers not involved as a team member in the anesthesia care to record the variations in strategies employed by anesthesia teams when ventilating by anesthesia mask in their patients presenting for procedures under general endotracheal tube anesthesia. Primarily the students on research rotation will follow anesthesia teams planning to perform general endotracheal tube anesthesia for anesthesia care wherein they will record percentage of successful anesthesia mask ventilations wherein anesthesia teams will be variably using one or more of the following strategies based on their personal preference and clinical experience with the strategies:<br>(a) bag ventilation, and/or<br>(b) mechanical ventilation, and/or<br>(c) no ventilation<br>plus<br>(a) one hand one provider on mask, and/or<br>(b) two hands one provider on mask, and/or<br>(c) more than two hands meaning more than one provider on mask, and/or<br>(d) bilateral jaw-thrust, and/or<br>(e) oral airway, and/or<br>(f) nasopharyngeal airway<br>plus<br>(a) one person doing everything<br>(b) two persons doing it together<br>(c) three or more persons doing it as team |
| **10.** | Describe how the results will be used/applied:<br>If successful anesthesia mask ventilations will be observed as more common with some strategies than with other strategies, then quality assurance and quality improvement discussions among our anesthesia teams may be planned while our results will be shared as preprint publications |

| 11. | Describe the participants for the project: |
|---|---|
| | All patients presenting for general endotracheal tube anesthesia at Harper University Hospital will be included so that their anesthesia mask ventilations can be observed by anesthesia researchers not involved as a team member in the anesthesia care to record the variations in strategies employed by anesthesia team during anesthesia mask ventilations in their patients and to quantify percentage of successful anesthesia mask ventilations as corresponding to strategies employed by our anesthesia teams. |

| | |
|---|---|
| **Reviewer Comments:** | Research: Knowledge gained from this study is not intended to be applicable to a larger population beyond the site of data collection or the specific participants/subjects studied, therefore the project is not intended to contribute to generalizable knowledge, thus the study does not meet the regulatory definition of research (45 CFR 46.102(i)).<br><br>Human Subjects/Participants: The study involves human participants via prospective interaction with participants through observation(45 CFR 46.102(e)). Note: Response to Section D #2.1 [illegible] |
| **Authorized IRB Reviewer Signature:** | Heather Park-May Digitally signed by Heather Park-May Date: 2023.05.17 15:25:01 -04'00' |

**2023 084**
